# LungAI: A Deep Learning Convolutional Neural Network for Automated Detection of COVID-19 from Posteroanterior Chest X-Rays

**DOI:** 10.1101/2020.12.19.20248530

**Authors:** Aryan Gulati

**Affiliations:** Delhi Public School R.K. Puram

**Keywords:** COVID19, Radiological Imaging, Machine Learning, Deep Learning, Convolutional Neural Networks, Artificial Intelligence, Detection

## Abstract

COVID-19 is an infectious disease caused by the Severe Acute Respiratory Syndrome Coronavirus 2 (SARS-CoV-2). As of December 2020, more than 72 million cases have been reported worldwide. The standard method of diagnosis is by Real-Time Reverse Transcription Polymerase Chain Reaction (rRT-PCR) from a Nasopharyngeal Swab. Currently, there is no vaccine or specific antiviral treatment for COVID-19. Due to rate of spreading of the disease manual detection among people is becoming more difficult because of a clear lack of testing capability. Thus there was need of a quick and reliable yet non-labour intensive detection technique. Considering that the virus predominantly appears in the form of a lung based abnormality I made use of Chest X-Rays as our primary mode of detection. For this detection system we made use of Posteroanterior (PA) Chest X-rays of people infected with Bacterial Pneumonia (2780 Images), Viral Pneumonia (1493 Images), Covid-19 (729 Images) as well as those of perfectly Healthy Individuals (1583 Images) procured from various Publicly Available Datasets and Radiological Societies. LungAI is a novel Convolutional Neural Network based on a Hybrid of the DarkNet and AlexNet architecture. The network was trained on 80% of the dataset with 20% kept for validation. The proposed Coronavirus Detection Model performed exceedingly well with an accuracy of 99.16%, along with a Sensitivity value of 99.31% and Specificity value of 99.14%. Thus LungAI has the potential to prove useful in managing the current Pandemic Situation by providing a reliable and fast alternative to Coronavirus Detection given strong results.

## II. Introduction

Emerging infectious diseases, such as those of the Severe Acute Respiratory Syndrome (SARS) have in the past and still continue to pose as a major threat to humanity. Yet despite various efforts and advancements in the field pertaining to infectious diseases little can be predicted and learnt beforehand about new and emerging diseases [1]. In early December 2019, initial cases of a then unknown form of Pneumonia were identified in Wuhan, China. By February 2020, over 80,000 laboratory confirmed cases had been recorded which led to the World Health Organization (WHO) declaring it as a Public Health Emergence of International Concern. Subsequently on 11th March 2020 the WHO declared COVID-19 as a pandemic [2].

**Fig 1.**
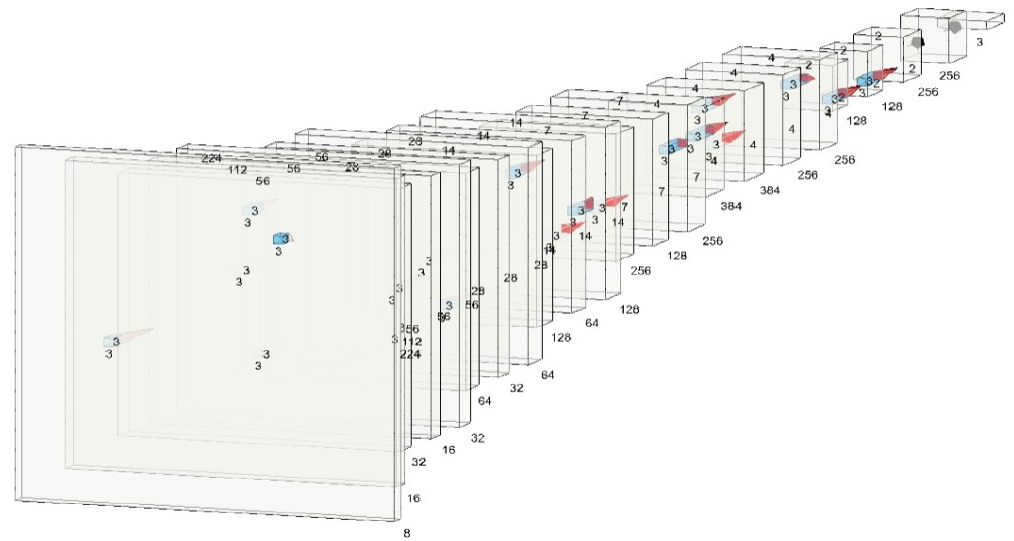
LungAI Convolutional Neural Network

Coronaviruses (CoVs) are clustered under the viral group that cause diseases in mammals and birds but while primarily found in bats, they rarely ever affected human beings. However, later due to various mutations the first coronavirus infected the nasal cavity of humans leading to symptoms resembling those of common cold. These viruses have a single strand RNA genome with a length ranging from 27 kilobases to 34 kilobases [3]. In the case of CoV infections in humans, they mostly involve affecting the upper respiratory tract and gastrointestinal tract with symptoms ranging from mild common cold like to severe cases of pneumonia and bronchitis [4].

The most recent of these coronaviruses is the Sars-CoV-2 virus which has affected millions of people around the world since the beginning of the year and has led to the world coming to a complete halt. Sequencing of the Sars-CoV-2 virus further shows that the virus genome shares 80% of its identity with that of Sars-CoV [5].

The pathology of COVID-19 also greatly resembles that of SARS with typical symptoms including fever, dry cough, shortness of breath and fatigue. Chest X-Ray image of afflicted patients show rapid progression of pneumonia and extensive consolidation in most patients. Interstitial and alveolar opacities mostly on periphery of lungs were observed largely involving both lungs. In some cases patients had pleural effusion. Features observed also included Ground Glass Opacity predominantly in the periphery of lungs [6], [7].

The speedy and exponential global spread of COVID-19 has lead to a strong urge for development of various techniques for its diagnosis [8],[9]. While much advancement has been made, the nucleic acid detection based approaches have become the go to choice for reliable detection of the Sars-CoV-2 virus. Most agencies have adopted the Polymerase Chain Reaction (PCR) route for detection of viruses, considering its relatively higher reliability. The Real Time PCR (RT-PCR) tests are today the basis on which communities are conducting their surveillance, prognosis of the prevailing situation and formulating Covid battling strategies. The question one cannot deny is that is the RT-PCR method reliable enough? What about the false-negative and false-positive results it is known to throw up. In view of the magnitude of testing required, is such accuracy acceptable and workable in the long run? The effect of false-negatives communicating the virus may lead to an exponential increase in cases which will finally defeat all measures taken to fight the pandemic. Moreover, the RT-PCR test’s dependence on use of gene primers may disallow it from detecting positive cases if the Sars-CoV-2 genome mutates. The virus is known to have variations in its viral RNA. Therefore, while as of date RT-PCR tests are the most viable, they have their limitations and its results need to be interpreted with caution. This apart, for common users these tests take anything upwards of 2 days which may lead to a delay in commencing isolation actions and treatment. [10]. Other major challenges in rapid detection are that test kits take longer duration for diagnosis of disease as well as longer time in production of probes, primers, and physical equipment. This has also lead to global shortage of testing kits in populations [11].

Rapid progress of medicine and its intertwining with various computational approaches over the last few years have lead to various developments. The progress Artificial Intelligence (AI) has made over the past few years has been unprecedented and development of Convolutional Neural Network (CNN) layers has allowed for significant gains in the ability to classify images and detect objects in a picture. One such field is Medical imaging, used to create images of the human body for medical procedures and diagnosis among others [12], [13]. Medical Imaging is already being widely used with Stanford University developing a Deep Learning based Neural Network that can classify over 2000 skin disease types including cancer and which successfully outperformed actual Dermatologists [14]. In a similar turn of events, there has also been advancement in the field of Computer Aided Diagnosis of Diabetic Retinopathy by making use of Eye Fundus Images [15], [16].

It was only a matter of time before Machine Learning applications found their way in detecting various lung abnormalities. Various forms of research have analyzed Lung Imaging in the form of either X-Rays or CT Scans as their baseline approaches for diagnosing certain illnesses. Many applications of Neural Networks are already showing better results than current techniques in Lung Cancer Detection with some having an accuracy of over 99% [17], [18]. There have also been applications of AI using CNNs in detection of other lung issues such as Tuberculosis, Pneumonia etc which have shown strong performances with accuracies in the high 90% [19] and thus showing a clear ability of Machine Learning approaches in diagnosing such diseases based on information procured from a series of lung images. This approach has also been applied to develop certain COVID19 detection protocols more of which has been talked about in the following section.

## III. Related Work

With the advent of Artificial Intelligence in medical imaging studies, Radiographic Chest X-Ray images have proven to be a valuable resource to form an alternative to the RT-PCR Tests. Some of the recent applications of Deep Learning in the field of COVID-19 detection include Ioannis D. Apostolopoulos & Tzani A. Mpesiana [20] who made use of Transfer Learning on State-Of-The-Art Pre-Trained models like VGG19, MobilenetV2, Inception, Xception etc of which the VGG19 achieved the highest accuracy in the Binary classifier of 98.75% as well as the highest accuracy in the multi class detection between COVID-19, Pneumonia and Normal of 93.48%. Their results were indicative of the superiority of VGG19 as compared to other models in terms of accuracy, sensitivity and specificity. Ozturk, T et al. [21] developed a DarkCovidNet CNN based on the DarkNet architecture which requires raw Chest X-ray images to return a diagnosis. The model produced a classification accuracy of 98.08% for binary classes (COVID-19 vs No-Findings) and 87.02% for multi-class cases (COVID-19 vs No-Findings vs Pneumonia). Elene Firmeza Ohata et al. [22] make use of Transfer Learning on CNNs trained on the ImageNet dataset as a feature extractor for X-Rays and then use these as an input to various machine learning algorithms like Support Vector Machines (SVM), K-Nearest Neighbour (KNN), Bayes, Random Forests, Multilayer Perceptron (MLP) etc. for final classification. In the first dataset (COVID19 vs Young Pneumonia Patients) the MobileNet-SVM architecture achieves the highest accuracy of 98.5% while on the second dataset (COVID-19 vs Pneumonia of all Ages) the DenseNet201-MLP model achieves an accuracy of 95.6%. Neha Gianchandani et al. [23] have developed an Ensemble Model of VGG16 and DenseNet201 and have made use of Transfer Learning to give a three-class classification (COVID19 vs Normal vs Pneumonia). The testing accuracy recorded was 99.21%. They also developed a similar ensemble of VGG16 and ResNet152V2 for binary classification (COVID19 Positive vs COVID19 Negative) which had an accuracy of 96.15%. Turkoglu [24] proposed a detection system called COVIDetectioNet which utilizes Transfer Learning on the AlexNet architecture along with Support Vector Machine as a classifier. The model was tested on a dataset of 6092 X-ray images comprising of Normal (Healthy), COVID19 and Pneumonia and achieved an accuracy of 99.18%. In another study [25] tested four pre trained architectures namely, AlexNet, DenseNet201, ResNet18 and SqueenzeNet. These were tested on a binary classification task (COVID19 Pneumonia vs Normal) and a three-class classification as well (COVID19 Pneumonia vs Normal vs Viral Pneumonia) post data augmentation of the Chest X-rays procured. In both instances the SqueezeNet model achieved the highest accuracy of 98.3%. In another similar study [26] a classifier based on the CheXNet architecture was developed and Transfer Learned on to detect a three-way classifier of COVID19, Normal and Viral Pneumonia X-ray images. Results indicated an accuracy of 97.8%.

**Fig. 2.**
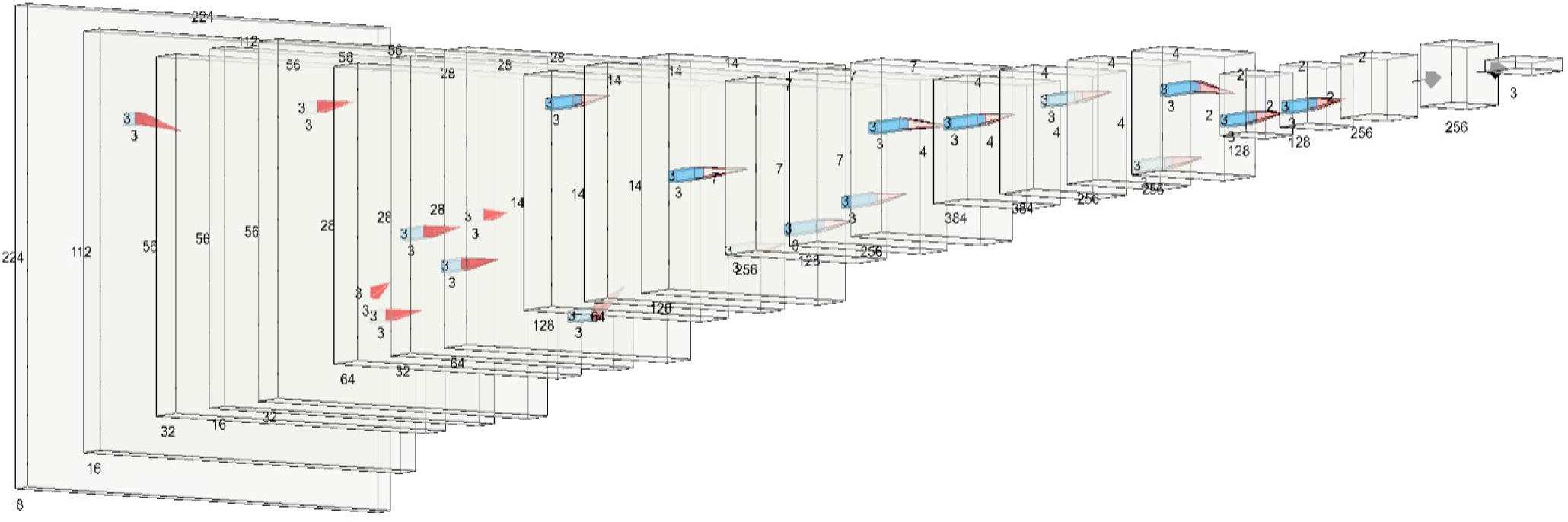
Architecture of Proposed LungAI Model

Yet despite the availability of a number of COVID19 detection tools there is still no sign of Artificial Intelligence approaches replacing the current nucleic-acid tests. Making use of this as the premise for my detection technique I developed LungAI, a novel Convolutional Neural Network that is based on the DarkNet-53 model architecture and the AlexNet model architecture. The main reason behind development of this Model was that most of the currently available solutions are forced to make use of Transfer Learning and Pre-Trained Models due to lack of enough training data on COVID19 to train a model from scratch as in most cases it would not yield results strong enough to be of any use. Thus, if a CNN developed from scratch could showcase results which either matched or bettered those of Transfer Learning with the low data and unbalanced datasets, given enough data in the future the CNN would surely outperform the current approaches. This would also ensure robustness of the developed CNN architecture which was and always has been the main aim. Another common problem faced by not just most AI solutions but also nucleic-acid testing is the high chance of giving False Negatives and False Positives due to a less competitive Sensitivity and Specificity score. Thus I also aimed my research towards developing an alternative that not only had a High Accuracy but also had the ability to limit results being False Negatives/Positives.

**TABLE I.**
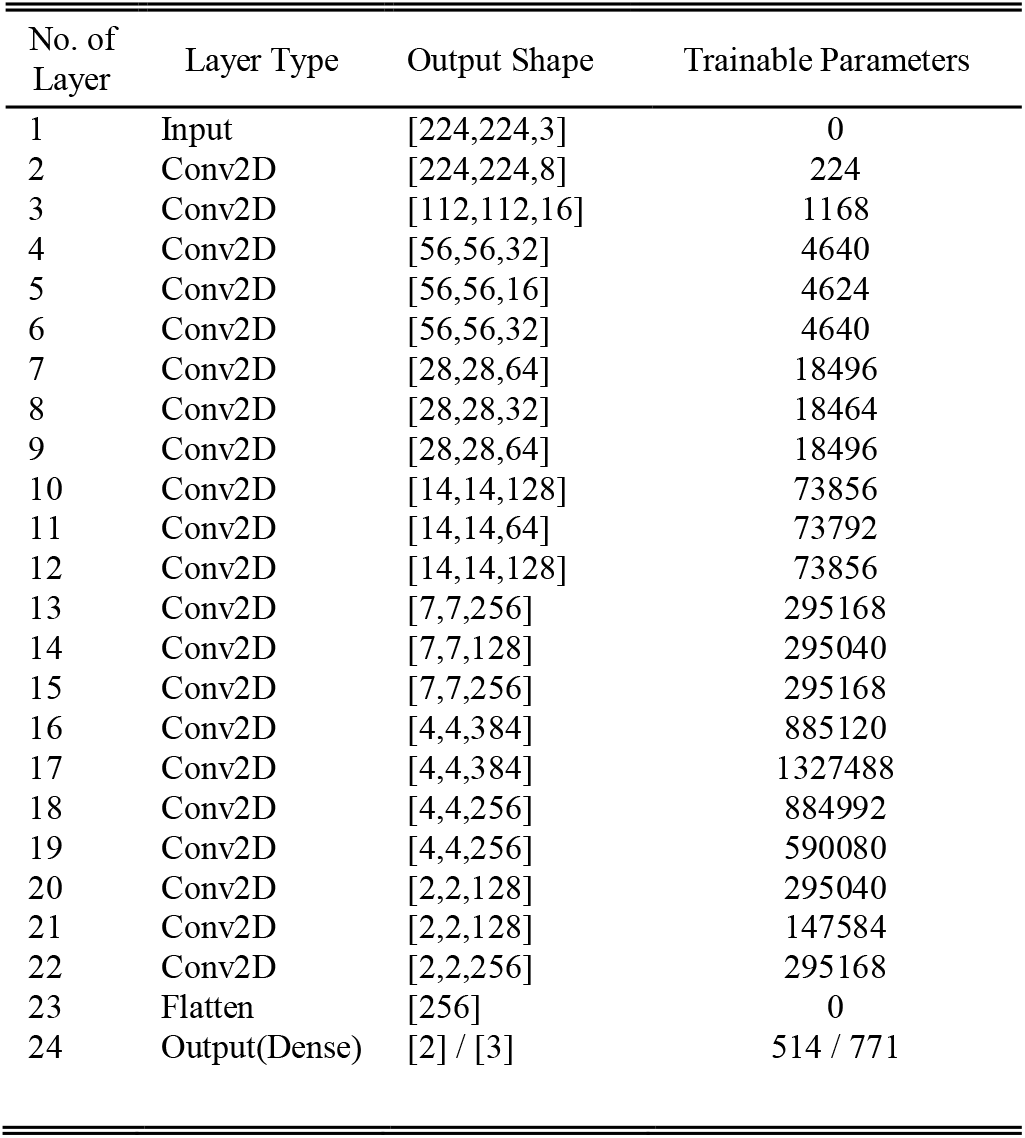
Layers and Parameters of LungAI (2 or 3 Way Classifications)

On testing and validation of LungAI, the network gave an Accuracy of 99.16% along with a Sensitivity of 99.31% and Specificity of 99.15% across a three-class classification task between COVID19, Normal and Pneumonia X-Rays. The model was also tested on 2 binary classifications tasks in which it yielded an Accuracy of 99.40%, Sensitivity of 99.31% and Specificity of 99.41% between COVID19 and Pneumonia as well as an Accuracy of 99.35%, Sensitivity of 100% and Specificity of 99.05% between COVID19 and Normal X-Rays. These results further go to show the robustness and high performance of the LungAI CNN Architecture in detecting presence of COVID19 among patients and will hopefully be of assistance in our fight against the pandemic and its effects.

In the following sections of this research paper I will describe in detail the methodology behind developing LungAI, the Lung Imaging Datasets that were used followed by the experimental results I obtained

**TABLE II.**
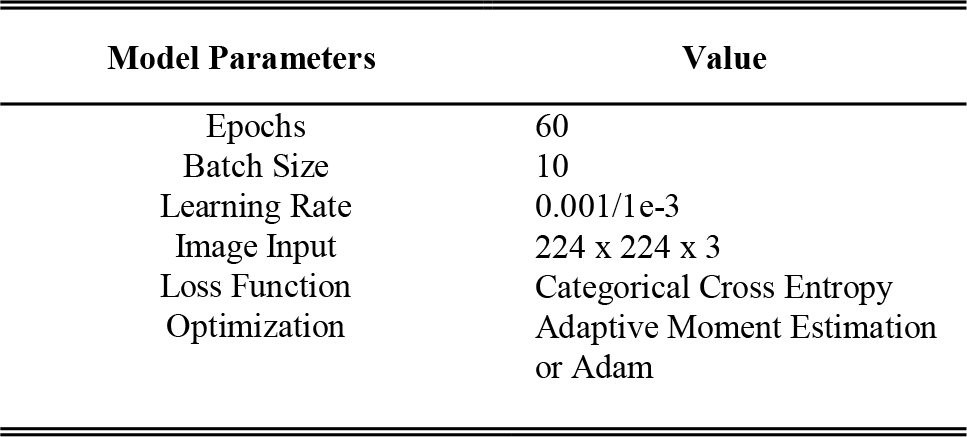
Associated Parameters for Model Training

## IV. Methods and Materials

### A. Proposed LungAI Convolutional Neural Network

Deep Learning refers to Machine Learning procedures conducted over Networks with multiple layers hence leading to the term ‘Deep’ which means depth of the Neural Networks defined by its layers. Convolutional Neural Networks (CNN) are a class of Deep Learning Networks used in tasks which require analysis of imaging data. While similar to Artificial Neural Networks (ANN), they explicitly assume that the input will be in the form of images.

Instead of developing a Deep Learning Architecture from scratch we made use of a more rational approach and thus analyzed existing CNN architectures before finally developing our own Novel Architecture. On further study of pre-existing architectures we narrowed it down to 2 models, namely the DarkNet53 which consists of 53 layers and AlexNet architecture which consists of 8 layers. For LungAI, we have used layers and filters with properties comparable to that of the above 2 models and have thus developed a Hybrid Architecture combining certain properties of the aforementioned DarkNet53 and AlexNet models.

Our proposed Deep Learning CNN consists of 21 2D Convolutional Layers, with each convolutional layer being followed by Batch Normalization and Rectified Linear Unit (ReLU) operations. Batch Normalization is used to make Neural Networks work faster while also maintaining stability as they normalize the output from previous activation layers by scaling and centering them. In certain cases Batch Normalization achieves the same accuracy with 14 times fewer training steps, and beats the original model by a significant margin [27]. The ReLU operations on the other hand ensure that there are no dying neurons by making sure that all neurons are not activated at the same time. Across the entire model we make use of Max Pooling layers for all pooling operations with 2D pooling size 2×2. Max Pooling layers as the name suggest select the highest values from filtered feature maps which ensure that the outputted feature map only contains prominent features while downsizing. It is also seen that Max Pooling layers work best in images that have a Dark Background which further prompted its usage in this case.

**Fig. 3.**
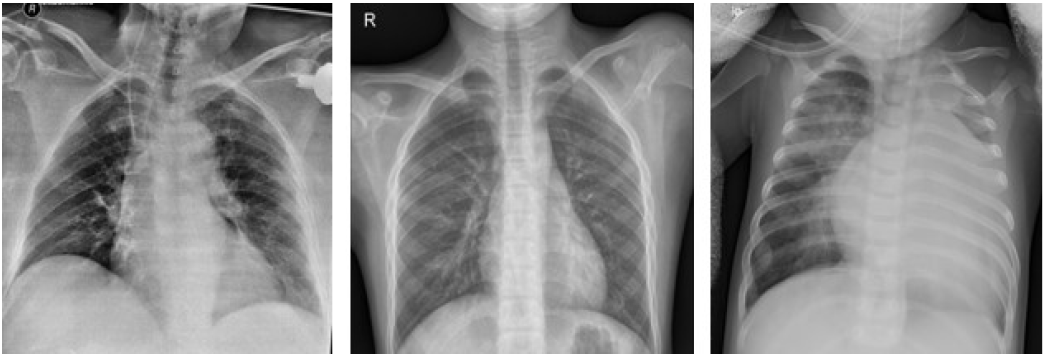
Chest X-Ray of (a) COVID19, (b) Normal & (c) Pneumonia affected patients

Lastly unlike most other Deep Learning CNNs LungAI’s architecture does not make use of Fully-Connected Layers (FC Layers) but rather makes use of Global Average Pooling Layer, followed by Flattening and then a final Output Layer with Softmax function. This change has multiple advantages as FC layers are prone to Overfitting and thus require use of Drop Layers to prevent the same which is not the case with Global Average Pooling. Another advantage comes in the form of Global Average Pooling being more native to convolutional structures than FC layers. Lastly, Global Average Pooling sums spatial information which makes it more robust than its alternative in this case [28].

Over the 21 Convolutional Layers, we use a varying number of filters. The first convolutional layer makes use of 8 filters, and post that the number of filters gradually increases to 16, 32, 64, 128, and 256 with the maximum number of filters being 384. All kernel sizes are maintained at 3×3 throughout the network. Irrespective of the number of classes (Binary or Three-Class), the model architecture remains the same with the only change being the output neurons in the Softmax layer. In case of the LungAI Binary classification task we set the Softmax function’s value as 2 for the COVID19-Pneumonia and COVID19-Normal detection. In case of the three-class classification the value of the Softmax is set to 3 for the COVID19-Normal-Pneumonia detection. The developed LungAI model consists of 5,609,907 trainable parameters. The model trained for 60 Epochs with a Batch Size (BS) of 10. The Learning Rate (LR) of during training of the model was set to 1e-3. The model makes use of a Softmax Activation Function along with Categorical Cross Entropy Loss Function and Adam Optimizer for updating the network weights.

**TABLE III.** Spread of Images across Disease Classes

| Image Class | Total Images | Training Set | Testing Set |
| --- | --- | --- | --- |
| COVID19 | 729 | 584 | 145 |
| Normal | 1583 | 1266 | 317 |
| Pneumonia | 4273 | 3418 | 855 |

### B. Data Collection

One of the major requirements for developing a successful AI tool for detection of COVID19 was availability of datasets of Chest X-Rays which were also verified. But due how recent these developments are there is still a lack of sufficient data and balanced datasets out there. I made use of JP Cohen’s openly available COVID19 dataset [29] which at the time of publishing contained 123 COVID19 X-Rays but has since been updated regularly with the most recent being 290 COVID19 Positive X-Rays. Apart from this I also made use of the Italian Society of Medical and Interventional Radiology’s (SIRM) public COVID19 database which consists of a mix of Chest X-Rays and CT Scans [30]. Lastly we also made use of the COVID-Net dataset which is an Open Source initiative taken up to provide and regularly update COVID19 data to further help in progressing detection based research of AI applications worldwide. The dataset currently contains 617 COVID positive X-Rays. Steps to replicate the dataset are also given here [31]. For the Normal and Pneumonia Chest X-Ray images we made use of the Kaggle Chest X-Ray dataset which consists of 5856 Chest X-Ray images spread out across 3 classes: Normal, Viral Pneumonia and Bacterial Pneumonia [32].

**TABLE IV.**
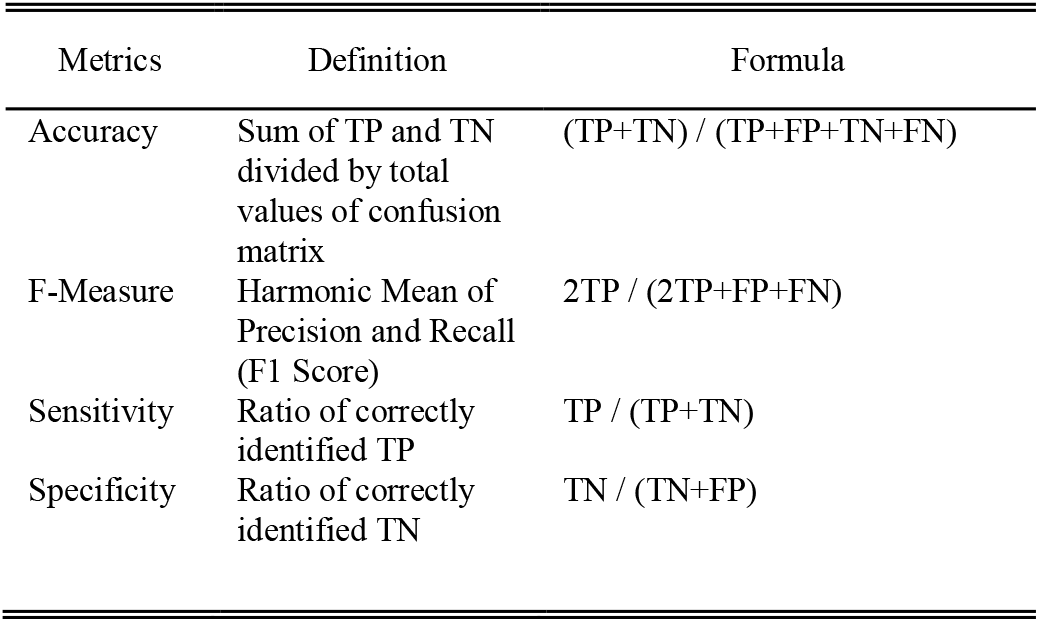
Evaluation Metrics

| Metrics | Definition | Formula |
| --- | --- | --- |
| Accuracy | Sum of TP and TN divided by total values of confusion matrix | $(TP+TN) / (TP+FP+TN+FN)$ |
| F-Measure | Harmonic Mean of Precision and Recall (F1 Score) | $2TP / (2TP+FP+FN)$ |
| Sensitivity | Ratio of correctly identified TP | $TP / (TP+TN)$ |
| Specificity | Ratio of correctly identified TN | $TN / (TN+FP)$ |

### C. Data Pre-Processing

Normalization refers to a method that changes the range of pixel intensities. Medical imaging plays a major role in various diagnostic procedures thus corrupted data could potentially lead to a misdiagnosis or worse. Yet sometimes due to noise or other factors medical machinery may give an inappropriate or incorrect description. To limit this issue, analyzed regions of interest in the image are often normalized [33].

In case of data pre-processing, the image files were further resized to 224 × 224 pixels. Apart from resizing to ensure compatibility with various programming packages the images were also made to swap colour channels from BGR(Blue, Green, Red) to RGB(Red, Green, Blue). I also made use of normalization to generate binary values which required converting all the data and labels to NumPy arrays while scaling the pixel intensities to the range of [0,255] due to there being 256 colour values (0-255). The images in the X-Ray dataset were normalized and then divided by 255 and mapped between 0 and 1 according to probability.

## V. Experimental Results and Discussion

### A. Training and Validation

The Model was trained over 60 epochs with a learning rate of 1e-3/0.001 along with Categorical Cross Entropy as the loss function. Graphs of LungAI’s training and validation loss and accuracy are given below. As can be observed from the Graphical Analysis of the CNNs loss function we see a significant increase in the loss value towards the beginning of the training procedure followed by a sharp decrease. This sudden and steep increase and decrease can be attributed to the unbalanced dataset where the numbers of samples of COVID19 are far lesser than its counterparts (Normal and Pneumonia). However further examination of the graph through the later stages shows rectification of this as by then the model has analyzed the complete dataset more than once and thus the crests and troughs in the graph are more subdued. This further also explains the commensurate sharp fall and rise in the validation accuracy at the same time for the CNN (Immediately Prior to Epoch 10).

**Fig. 4.**
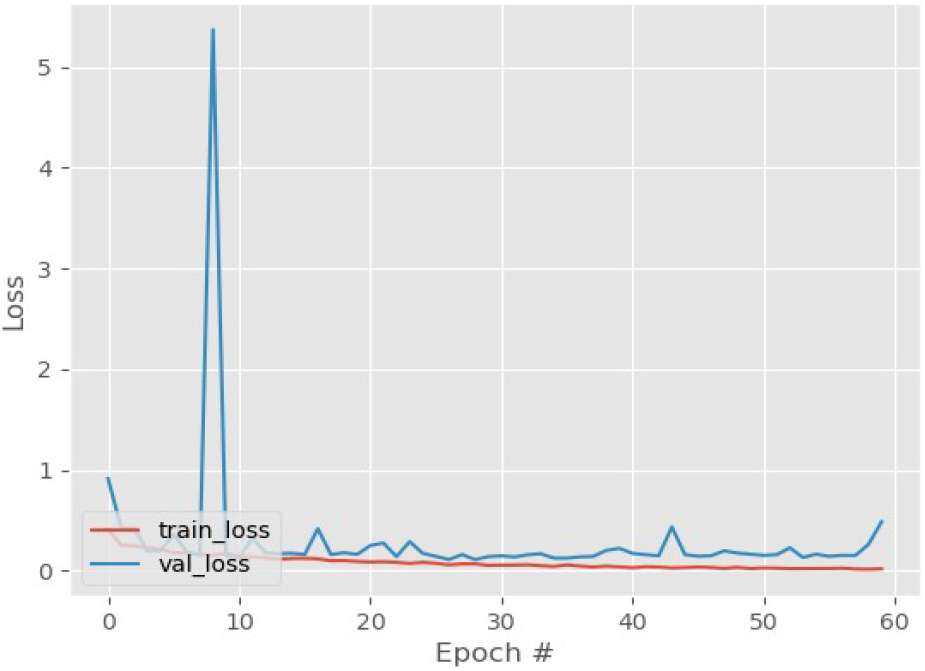
LungAI Training & Validation Loss

**Fig. 5.**
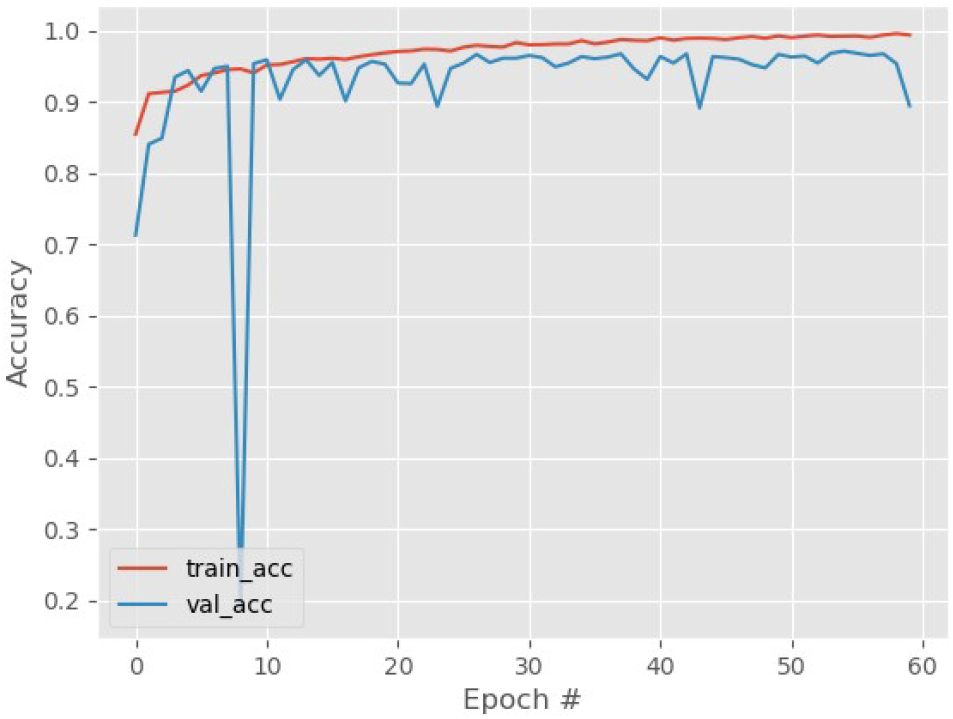
LungAI Training & Validation Accuracy

### B. Evaluation Metrics

Specific metrics for evaluation were recorded as follows: (a) Correctly identified cases (True Positives, TP), (b) Incorrectly classified cases (False Negatives, FN), (c) Correctly identified healthy cases (True Negatives, TN), and (d), Incorrectly classified healthy cases (False Positives, FP). Please note that TP refers to the correctly predicted Covid-19 cases, FP refers to Normal or Pneumonia cases classified as COVID19, TN refers to Normal or Pneumonia cases classified as Non-COVID19, while FN refers to COVID19 cases classified as Normal or as Pneumonia figures will be processed as images.

We performed experiments on validation test sets over 3 types of data. The first dataset was a three way classification between COVID19, Normal and Pneumonia while the other 2 datasets were for a binary classification between COVID19, Normal and COVID19, Pneumonia. The model was evaluated on the basis of Accuracy, Sensitivity, Specificity and F1 score. This was done so as to give equal importance to other factors affecting the networks COVID19 detecting ability including outputting of false positives and false negatives.

### C. COVID19 vs Normal vs Pneumonia (3 Way Classification)

**Fig. 6.**
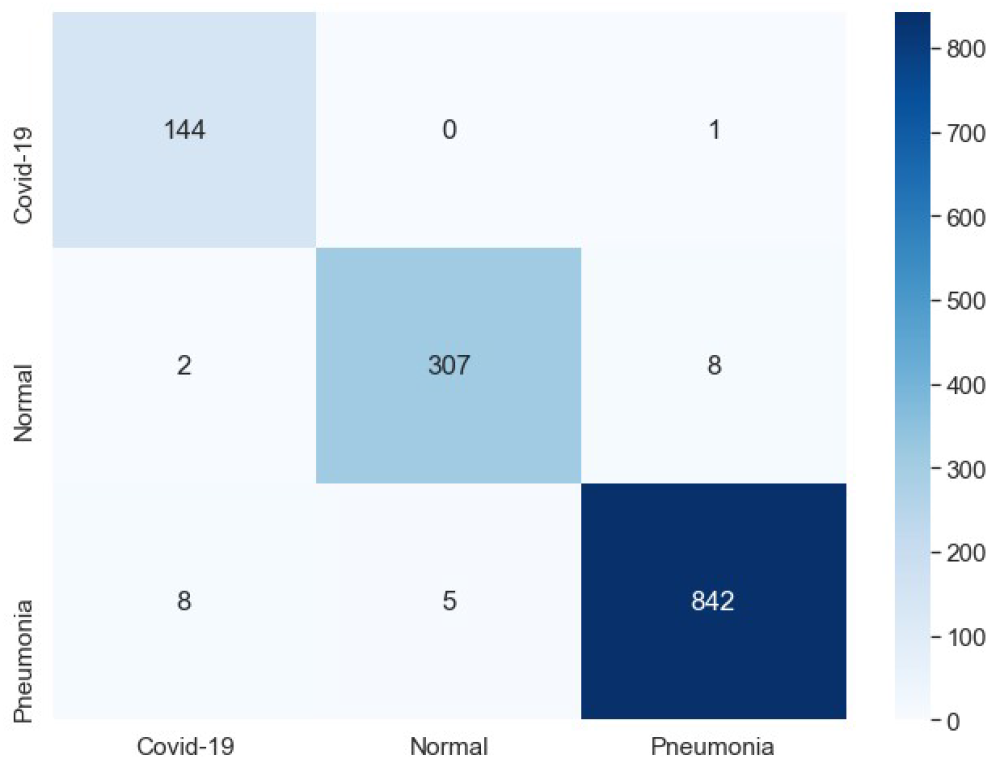
Confusion Matrix (COVID19, Normal & Pneumonia)

**TABLE V.** Model Performance Metrics (COVID19, Normal & Pneumonia)

| TABLE V<br>MODEL PERFORMANCE METRICS (COVID19, NORMAL & PNEUMONIA) |  |
| --- | --- |
| COVID19, Normal & Pneumonia) |  |
| Accuracy | 99.16% |
| F1 Score | 99.32% |
| Sensitivity | 99.31% |
| Specificity | 99.14% |

### D. COVID19 vs Pneumonia (2 Way Classification)

**Fig. 7.**
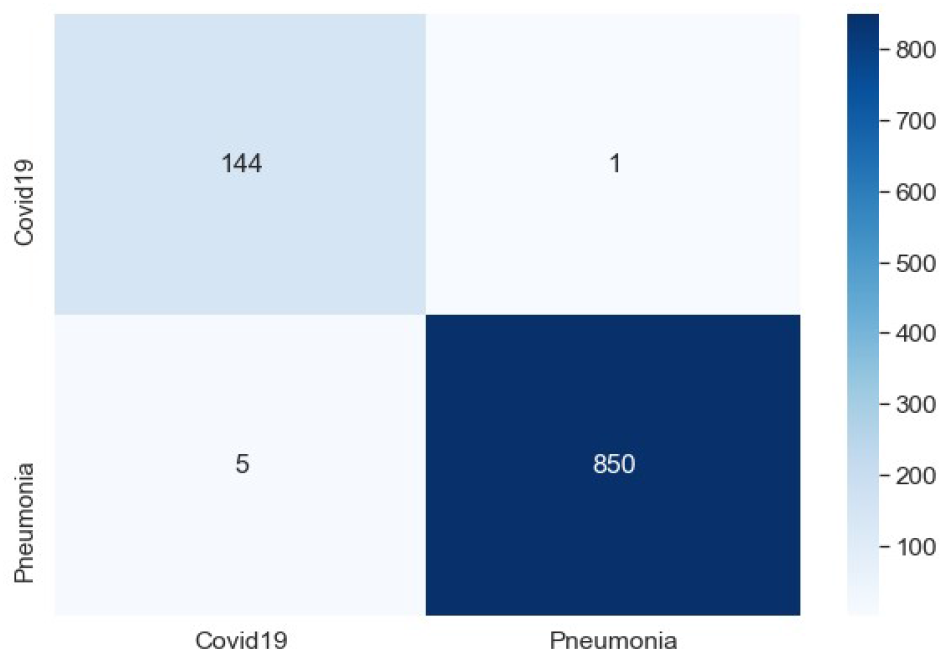
Confusion Matrix (COVID19 & Pneumonia)

**TABLE VI.** Model Performance Metrics (COVID19 & Pneumonia)

| TABLE VI<br>MODEL PERFORMANCE METRICS (COVID19 & PNEUMONIA) |  |
| --- | --- |
| COVID19 & Pneumonia) |  |
| Accuracy | 99.40% |
| F1 Score | 97.96% |
| Sensitivity | 99.32% |
| Specificity | 99.41% |

### D. COVID19 vs Normal (2 Way Classification)

**Fig. 8.**
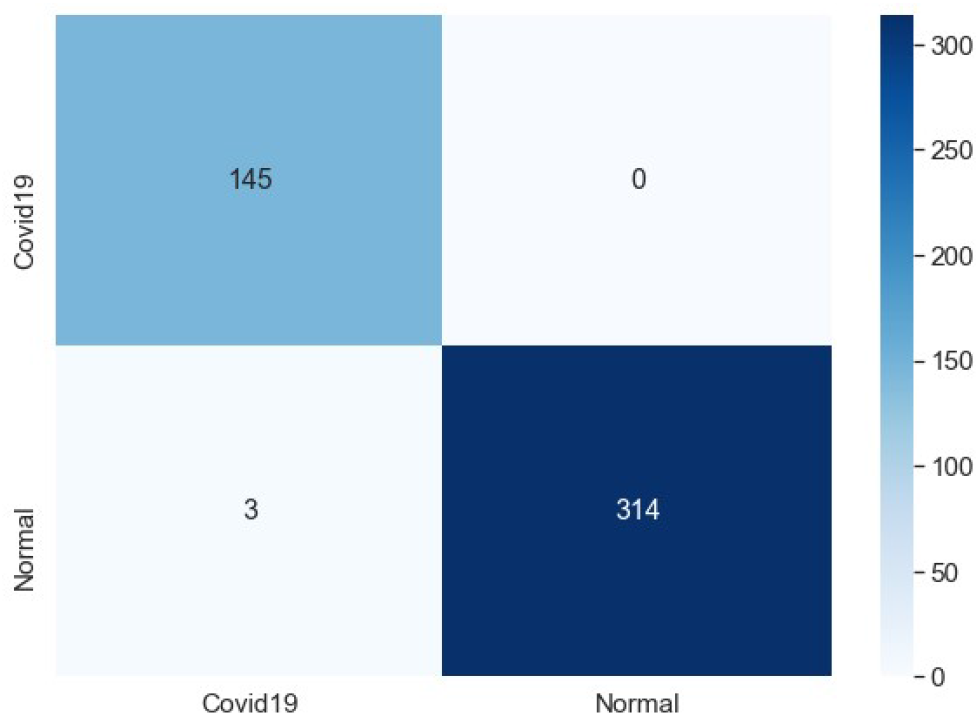
Confusion Matrix (COVID19 & Normal)

**TABLE VII.** Model Performance Metrics (COVID19 & Normal)

| TABLE VII<br>MODEL PERFORMANCE METRICS (COVID19 & NORMAL) |  |
| --- | --- |
| COVID19 & Pneumonia) |  |
| Accuracy | 99.35% |
| F1 Score | 98.97% |
| Sensitivity | 100.00% |
| Specificity | 99.05% |

## VI. Conclusion

Considering the potential of Artificial Intelligence, its utilization towards mass COVID19 surveillance is but imperative. Various efforts towards evolving algorithms for COVID detection have been mired by lack of data and inadequate Sensitivity and Specificity scores which disallows these programs from either getting the required accuracy. However, this void can be overcome by incorporating the LungAI Convolutional Neural Network architecture achieving an accuracy of over 99% along with an extremely degree of Sensitivity. The simplicity of use, speed and high reliability of this application along with the emergent requirement of developing an alternate mass surveillance system makes this architecture promising for further refinement. If adapted, this workable model may form the foundation of Anti-COVID strategies being developed by communities across the globe.

## Data Availability

The datasets analyzed during the current study are available in various public and open repositories.

https://github.com/ieee8023/covid-chestxray-dataset

https://www.sirm.org/en/category/articles/covid-19-database/

https://www.kaggle.com/paultimothymooney/chest-xray-pneumonia

